# Etomidate Versus Ketamine for Emergency Intubation in Critically Ill Patients: An Updated Meta-Analysis and Systematic Review

**DOI:** 10.64898/2026.02.27.26347260

**Authors:** Vítor H. Andriazzi, Renato P. Curcio, Michael A. R. A. Novais, Bernardo L. G. Fernandes, Giovana C. Rosa, Juan G. S. Vasconcelos, Juliano N. Quineper

## Abstract

**Objective:** To compare the efficacy and safety of etomidate versus ketamine as induction agents for rapid sequence intubation in critically ill adults, focusing on 28-day mortality and post-intubation hypotension.

**Data Sources:** PubMed, Embase, and the Cochrane Library were systematically searched from inception to January 2026. Reference lists of included studies were also manually screened.

**Study Selection:** We included randomized controlled trials (RCTs) comparing single-dose intravenous ketamine versus etomidate for emergency rapid sequence intubation in critically ill adults (≥ 18 years) in non-operating room settings (e.g., intensive care unit or emergency department).

**Data Extraction:** Two investigators independently screened records, extracted data using a standardized form and assessed the risk of bias using the RoB 2 tool. The certainty of evidence was evaluated using the GRADE framework.

**Data Synthesis:** Six RCTs comprising 4,108 patients (2,046 assigned to ketamine and 2,062 to etomidate) were included. The pooled analysis showed no statistically significant difference in 28-day mortality between the ketamine and etomidate groups (39.0% vs. 40.3%; relative risk [RR] 0.96; 95% CI, 0.89–1.03; p=0.29; I²=11%). In a prespecified subgroup analysis of patients with sepsis (n=1,546), mortality also did not differ significantly (RR 0.94; 95% CI, 0.86–1.03). However, ketamine was associated with a statistically significant increase in the incidence of post-intubation hypotension (14.2% vs. 11.3%; RR 1.25; 95% CI, 1.01–1.53; p=0.04; I²=0%). No significant differences were observed regarding peri-intubation cardiac arrest, first-attempt intubation success, or ventilator- and intensive care unit-free days.

**Conclusions:** There is no statistical difference in 28-day mortality between etomidate and ketamine for emergency intubation in critically ill adults, including those with sepsis. The higher incidence of post-intubation hypotension with ketamine suggests etomidate presents a more favorable hemodynamic safety profile in this setting.

**Key points:** *Question:* Does the choice between etomidate and ketamine for emergency intubation in critically ill patients impact 28-day mortality?

*Findings:* In this systematic review and meta-analysis of randomized controlled trials, there was no statistically significant difference in 28-day mortality between patients induced with ketamine (39.0%) and those induced with etomidate (40.3%).

*Meaning:* The use of etomidate versus ketamine for rapid sequence intubation does not alter 28-day mortality, indicating that the choice of induction agent should be individualized.

## 1. Introduction

Rapid sequence intubation (RSI) is the standard of care for securing the airway in critically ill patients, a high-risk procedure associated with significant rates of life-threatening complications, including severe hypotension and cardiac arrest(1,2). Selecting the induction agent is critical to mitigating these risks. Etomidate and ketamine are the two most commonly used agents in this setting due to their favorable pharmacokinetic profiles and rapid onset of action(3,4).

Etomidate has traditionally been favored for its hemodynamic stability(5). However, it is well-established that even a single bolus dose inhibits the enzyme 11β-hydroxylase, causing reversible adrenal suppression(6). This “pharmacologic adrenalectomy” has raised persistent concerns regarding its safety in patients with sepsis, potentially impairing the physiological stress response and increasing mortality(7). Conversely, ketamine is a dissociative anesthetic with sympathomimetic properties that theoretically maintain blood pressure through catecholamine release(8). Consequently, ketamine has been increasingly advocated for use in patients with shock(9).

Despite these theoretical advantages, the clinical superiority of one agent over the other remains controversial. While etomidate causes adrenal inhibition, it is unclear if this translates to increased mortality or vasopressor requirements(10–12). On the other hand, the hemodynamic safety of ketamine has been challenged; recent evidence suggests it may precipitate cardiovascular collapse in catecholamine-depleted patients, such as those with prolonged septic shock(13,14). Current clinical practice guidelines do not recommend one agent over the other due to conflicting evidence(15).

Prior systematic reviews and meta-analyses have attempted to resolve this debate but have been limited by the inclusion of observational studies, significant heterogeneity, or small sample sizes(11,12,16). Recently, large-scale randomized controlled trials (RCTs) have been published, contributing substantial new data regarding mortality and peri-intubation hemodynamic stability(17,18). We therefore conducted a systematic review and meta-analysis of RCTs, including these most recent trials, to compare the efficacy and safety of etomidate versus ketamine in critically ill adults, with a specific focus on 28-day mortality and post-induction hypotension.

## 2. Methods

This systematic review follows the Cochrane Handbook for Systematic Reviews of Interventions(19) and complies with the Preferred Reporting Items for Systematic Reviews and Meta-Analyses (PRISMA) guidelines(20); the protocol was prospectively registered in PROSPERO (CRD420261279883).

### 2.1 Eligibility criteria

Studies were deemed eligible for inclusion if they met the following criteria: (a) study design consisting of randomized controlled trials (RCTs) conducted in non-operating room settings, such as the emergency department (ED) or intensive care unit (ICU); (b) adult patients (aged ≥ 18 years) with critical illness requiring emergency RSI; (c) direct comparison of single-dose intravenous ketamine versus etomidate as the primary induction agent; and (d) reporting clinical outcomes of interest. This restriction to non-operating room settings was applied specifically to exclude elective surgical populations. Unlike stable surgical patients, critically ill adults frequently present with catecholamine depletion and hemodynamic instability(14). Excluding elective populations ensures the analysis specifically addresses the safety profile of ketamine in patients where its negative inotropic properties might precipitate cardiovascular collapse, rather than its indirect sympathomimetic effects seen in healthy physiology(21–23). To ensure the analysis focused on high-quality evidence and this specific physiologic profile, we applied the following exclusion criteria: (a) observational studies; (b) trials involving pregnant patients; (c) prisoners; and (d) patients in cardiac arrest prior to intubation.

### 2.2 Search strategy and data extraction

We systematically searched PubMed, Embase, and the Cochrane Library from their inception through January 2026, using a combination of controlled vocabulary and keywords relevant to ’etomidate’, ’ketamine’, ’rapid sequence intubation’, and ’critical illness’. To ensure comprehensive coverage, we also manually screened the reference lists of all included studies and relevant systematic reviews to identify potential trials not captured by the electronic search. Following the search, two investigators independently screened titles and abstracts for eligibility and extracted data from the included studies using a standardized form, strictly adhering to the pre-established inclusion criteria; any discrepancies were resolved through discussion or adjudication by a third reviewer.

### 2.3 Endpoints and sensitivity analysis

Outcomes of interest included 28-day mortality, periprocedural cardiac arrest, first-attempt intubation success, post-intubation hypotension, ICU-free days, and mechanical ventilator-free days. We acknowledge that endpoint definitions varied among the included trials; for instance, cardiac arrest definitions differed slightly regarding the peri-intubation time window, and hypotension was defined by some authors as a percentage decrease from baseline (e.g., >20% drop), while others used absolute systolic blood pressure thresholds ranging from <90 mmHg to <80 mmHg(18,24,25). We performed a prespecified subgroup analysis restricted to patients with sepsis to evaluate safety in this high-risk population. Additionally, to assess the robustness of our findings and address potential publication bias given the limited number of trials for a funnel plot(19), we conducted a sensitivity analysis incorporating data from an unpublished quasi-randomized trial identified in gray literature. However, these studies were excluded from the primary analysis and GRADE certainty assessment; this decision was made *a priori*, prior to data synthesis, to ensure methodological rigor independent of the direction of the effect.

### 2.4 Quality assessment

Two investigators independently assessed the risk of bias for each included trial using the Revised Cochrane Risk of Bias tool for randomized trials (RoB 2)(26). We evaluated bias across five specific domains: randomization process, deviations from intended interventions, missing outcome data, measurement of the outcome, and selection of the reported result. The overall risk of bias was categorized as ’low risk,’ ’some concerns,’ or ’high risk.’ Additionally, the certainty of the evidence for primary and secondary outcomes was evaluated using the Grading of Recommendations Assessment, Development, and Evaluation (GRADE) framework(27). Any discrepancies in the quality assessment were resolved through consensus.

### 2.5 Statistical analysis

For binary outcomes, we calculated risk ratios (RRs) with 95% confidence intervals (CIs), while mean differences (MDs) were used for continuous outcomes. Since continuous variables in the included trials were predominantly reported as medians with interquartile ranges (IQRs), we estimated the sample means and standard deviations using the method described by Wan et al(28). Statistical heterogeneity was evaluated using the Cochrane Q test and the I2 statistic(19); heterogeneity was considered significant if the Q test p-value was < 0.10 or I2 > 50%. We pooled data using the Restricted Maximum Likelihood random-effects model to account for potential between-study variance. All statistical analyses were performed using R software, version 4.5.2 (R Foundation for Statistical Computing, Vienna, Austria)(29).

### 2.6 Data availability

The template data collection forms, data extracted from included studies, data used for all analyses, and analytic code generated during this systematic review are not publicly available, but can be obtained from the corresponding author upon request.

## 3. Results

### 3.1 Study selection and baseline characteristics

The initial electronic search identified a total of 1680 records. After removing duplicates and clearly ineligible studies, the 26 remaining articles were fully reviewed for eligibility. Ultimately, 6 randomized controlled trials(17,18,24,25,30,31) were included in the quantitative synthesis. The detailed screening process is illustrated in the PRISMA flow diagram (Figure 1).

**Figure 1.**
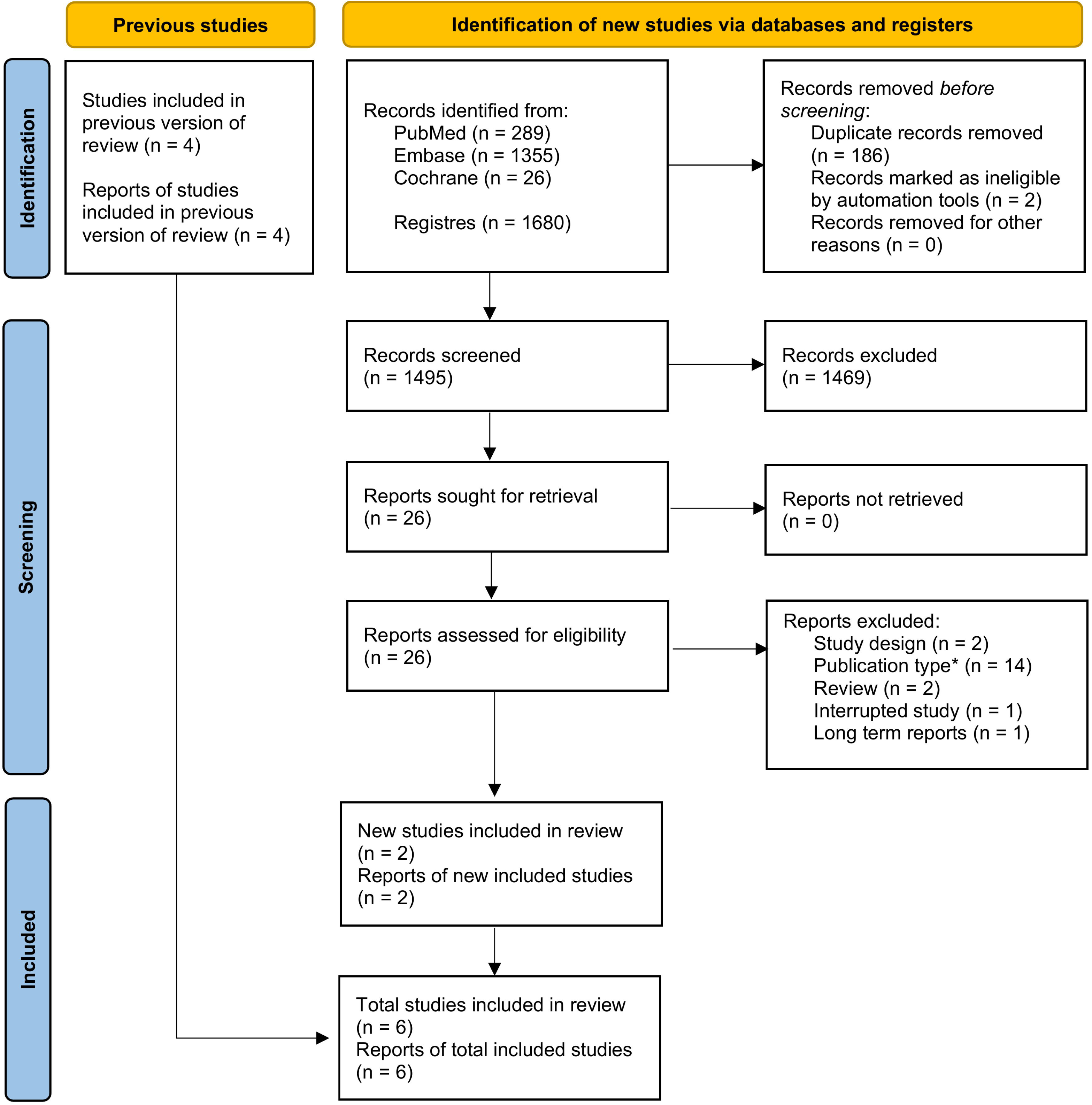
PRISMA flow diagram detailing the study selection process. *Including notes (1), editorials (1), protocols (2), letters (5), commentaries (2), interim analysis (1) and duplicated trial registries (2). Adapted from: Page MJ, et al. The PRISMA 2020 statement: an updated guideline for reporting systematic reviews. BMJ. 2021. (20)"

A total of 4,108 randomized patients were included in the quantitative synthesis, with 2,046 (49.8%) assigned to the ketamine group and 2,062 (50.2%) to the etomidate group. The recent multicenter trial contributed most of the pooled population (n = 2,365)(18). The included trials exhibited significant clinical heterogeneity regarding the prevalence of sepsis: while some studies exclusively enrolled patients with sepsis(25,30), this condition was present in less than 20% of the cohorts in others(24,31). Regarding the intervention, dosing protocols varied between fixed weight-based doses and weight-based ranges across trials, with ketamine doses ranging from 1 to 2 mg/kg and etomidate doses ranging from 0.2 to 0.3 mg/kg. Despite these variations, baseline demographics were generally well-balanced between the study arms across all trials (Table 1).

**Table 1.** Baseline characteristics of included studies.

| Study | Source | Design | Patients<br>Ket/Eto | Sepsis, %<br>Ket/Eto | Male, %<br>Ket/Eto | Age<sup>d</sup>, y<br>Ket/Eto | SBP<sup>d</sup>, %<br>Ket/Eto | HTN, %<br>Ket/Eto | DBT, %<br>Ket/Eto | ARF, %<br>Ket/Eto | Ketamine<br>(mg/kg) | Etomidate<br>(mg/kg) |
| --- | --- | --- | --- | --- | --- | --- | --- | --- | --- | --- | --- | --- |
| Jabre 2009 (32) | Journal's<br>article | MC RCT | 235/234 | 15/18 | 57/63 | 59/57 | 128/132 | 34/33 | 13/14 | 17/16 | 2 | 0.3 |
| Matchett 2022<br>(17) | Journal's<br>article | SC RCT | 395/396 | 50/52 | 62/61 | 55/56 | 120/121 | NA | NA | 48/44 | 1 or 2 | 0.2 or 0.3 |
| Knack 2023<br>(24) | Journal's<br>article | SC RCT | 70/73 | 14/26 | 60/67 | 50/49 | 139/140 | 29/33 | NA | 17/27 | 2 | 0.3 |
| Srivilaithon<br>2023 (25) | Journal's<br>article | SC RCT | 130/130 | 100 | 59/59 | 71/73 | 118/113 | 59/62 | 45/37 | 40/42 | 1 or 2 | 0.2 or 0.3 |
| Agarwal 2025<br>(31) | Journal's<br>article | SC RCT | 40/40 | 100 | 58/60 | 71/73 | 118/113 | 58/63 | 45/38 | 40/43 | 1 or 2 | 0.2 or 0.3 |
| Casey 2025 (18) | Journal's<br>article | MC RCT | 1176/1189 | 46/48 | 58/59 | 60/60 | 115/114<sup>b</sup> | 46/45 | NA | 58/57<sup>a</sup> | 1, 1.5 or 2 | 0.2, 0.25 or<br>0.3 |

<sup>a</sup>reported as acute respiratory condition; <sup>b</sup>lowest pre-intubation systolic blood pressure registered; <sup>c</sup>significant statistical difference between ketamine and etomidate group; <sup>d</sup>mean or median; ARF: acute respiratory failure; DBT: Diabetes; Eto: Etomidate; HTN: hypertension; Ket: ketamine; NA: not available; MC: Multicenter; RCT: randomized controlled trial; SBP: pre intubation systolic blood pressure; SC: single-centre

### 3.2 Pooled analysis of all studies

Regarding the primary outcome, the pooled analysis of six trials showed no statistically significant difference in 28-day mortality between the ketamine (n = 2,046) and etomidate (n = 2,062) groups (39% vs. 40.3%; Risk Ratio [RR] 0.96; 95% Confidence Interval [CI], 0.89 to 1.03; P = 0.29). Heterogeneity for this outcome was low (I2 = 11%). (Figure 2, panel A)

**Figure 2.**
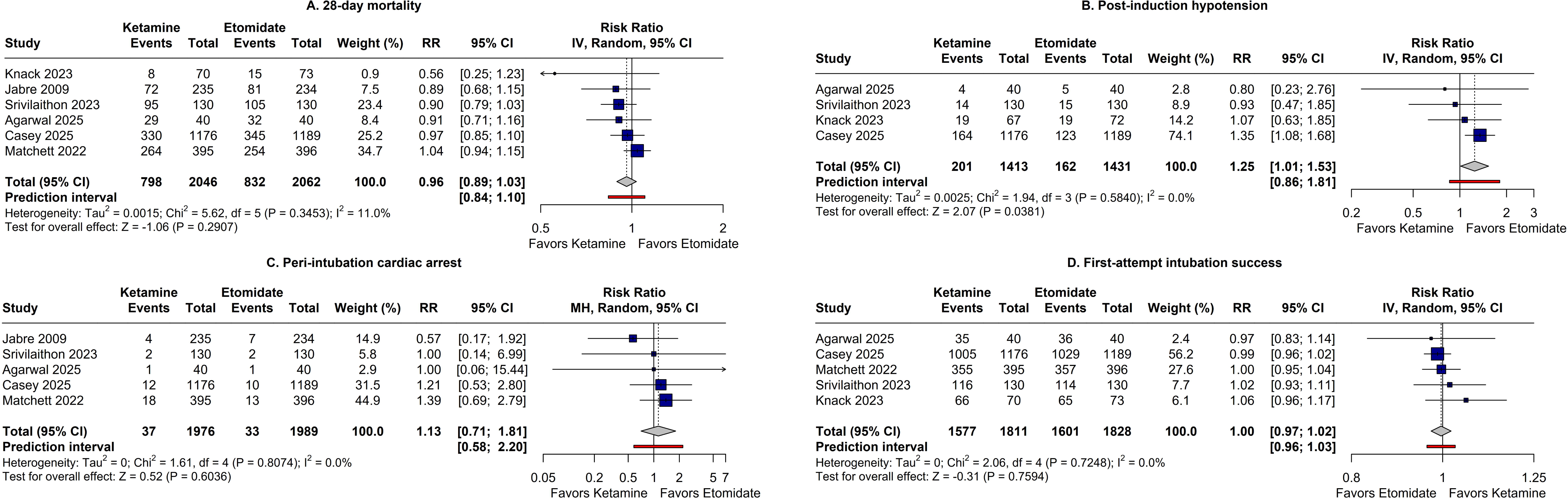
Forest plots comparing primary and secondary outcomes of rapid sequence intubation using ketamine versus etomidate. The panels illustrate the pooled effect estimates for (A) 28-day all-cause mortality, (B) post-intubation hypotension, (C) peri-intubation cardiac arrest, and (D) first-attempt intubation success. CI = Confidence Interval; IV = Inverse Variance; MH = Mantel-Haenszel; RR = Risk Ratio.

Regarding secondary outcomes, ketamine was associated with a statistically significant increase in the risk of post-intubation hypotension (14.2% vs. 11.3%; RR 1.25; 95% CI, 1.01 to 1.53; P = 0.04), with no observed heterogeneity (I2 = 0%). However, this hemodynamic difference did not translate into a significant difference in the incidence of peri-intubation cardiac arrest (1.9% vs. 1.7%; RR 1.13; 95% CI, 0.71 to 1.81; P = 0.60; I2 = 0%). Procedural efficacy, measured by first-attempt intubation success, was similar between groups (87.1% vs. 87.6%; RR 1.00; 95% CI, 0.97 to 1.02; P = 0.76). (Figure 2, panels B to D)

Furthermore, no significant differences were observed in resource utilization outcomes, including ICU-free days (Mean Difference [MD] -0.3 days; 95% CI, -1.5 to 0.9) and ventilator-free days (MD -0.3 days; 95% CI, -0.9 to 0.3). (Figure 3)

**Figure 3.**
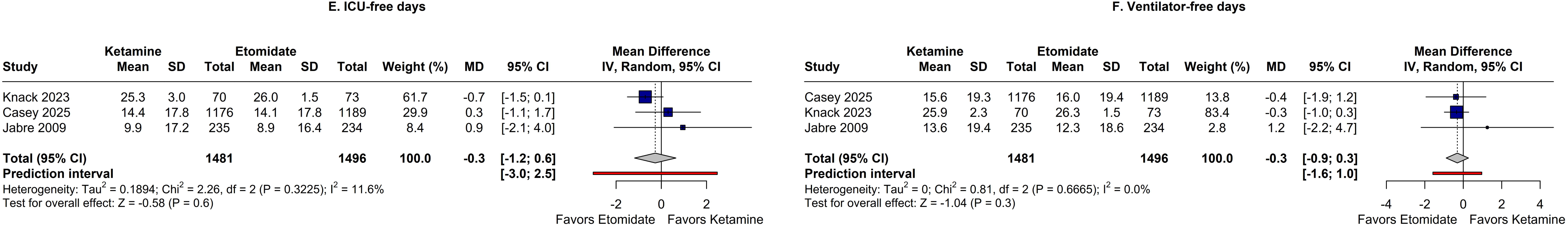
Forest plots comparing hospital resource utilization outcomes. The panels illustrate the pooled effect estimates for (E) ICU-free days and (F) ventilator-free days. CI = Confidence Interval; ICU = Intensive Care Unit; IV = Inverse Variance; MD = Mean Difference.

### 3.3 Subanalyses in selected populations

We performed a prespecified subgroup analysis to evaluate 28-day mortality stratified by the presence of sepsis. In the septic patient subgroup (n = 1,546), pooled data from five trials showed no significant difference in mortality between ketamine (46.0%) and etomidate (47.0%), with a risk ratio of 0.94 (95% CI, 0.86 to 1.03; P = 0.20) and no heterogeneity (I2 = 0%). Similarly, in the non-septic patient subgroup (n = 1,101), mortality rates were comparable between ketamine (34.9%) and etomidate (36.4%) (RR 0.89; 95% CI, 0.75 to 1.06; P = 0.20; I2 = 0%). The test for subgroup differences indicated no statistically significant interaction between sepsis status and the treatment effect (P = 0.60). (Figure 4)

**Figure 4.**
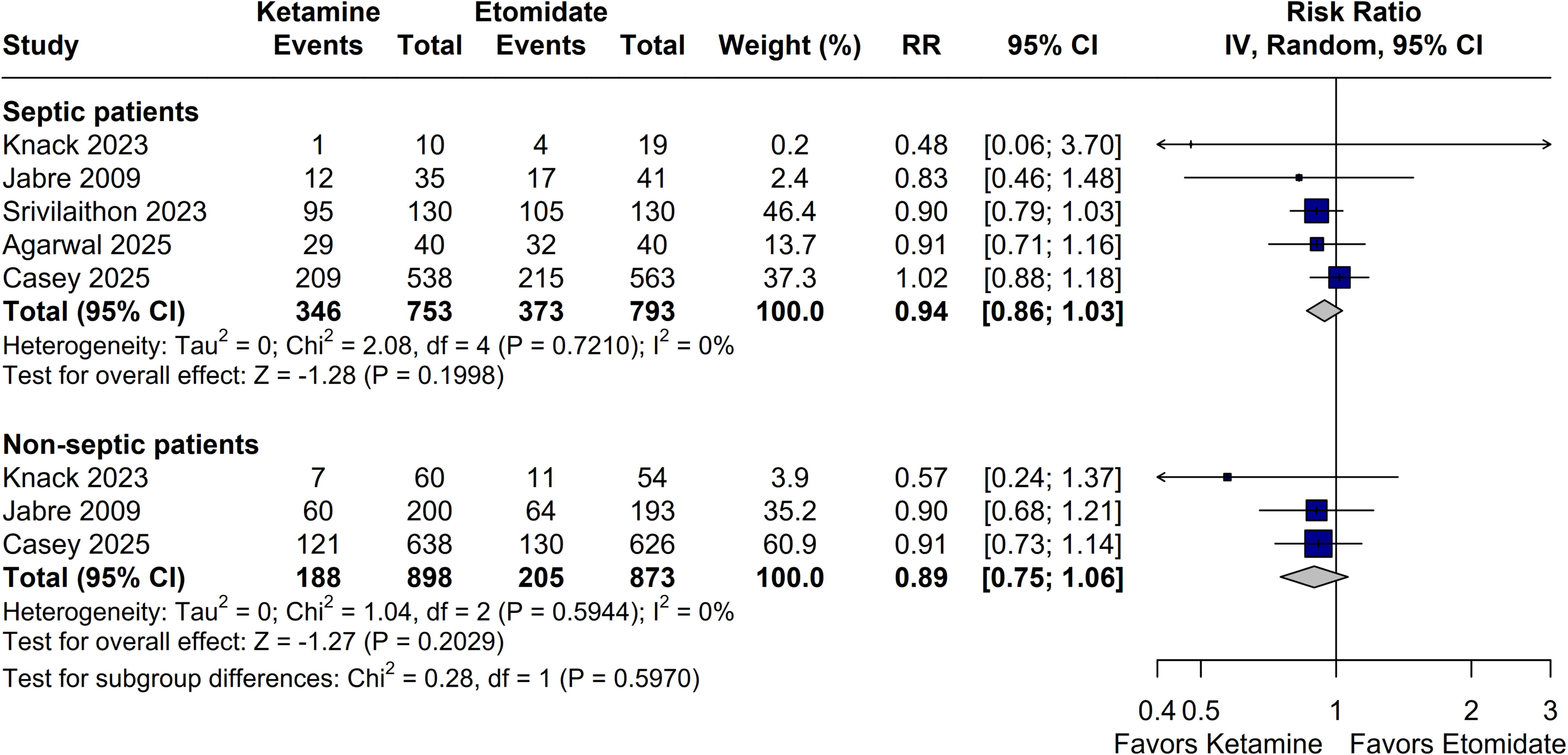
Subgroup analysis of 28-day mortality stratified by the presence of sepsis. Forest plots comparing the pooled effect estimates of rapid sequence intubation using ketamine versus etomidate in septic and non-septic patients. CI = Confidence Interval; IV = Inverse Variance; RR = Risk Ratio.

A sensitivity analysis was conducted for the outcome of post-intubation hypotension to assess the impact of including the Powers et al. trial (ClinicalTrials.gov NCT03545503, unpublished data) (eFigure 8). The inclusion of this study (n = 398) did not alter the direction or statistical significance of the primary finding. The pooled analysis of all five studies (n = 3,242) confirmed that ketamine was associated with a significantly higher risk of post-intubation hypotension compared to etomidate (15.7% vs. 12.2%; RR 1.29; 95% CI, 1.08 to 1.53; P = 0.004). Heterogeneity remained at 0% with the inclusion of this trial.

### 3.4 Quality assessment

Assessment using the RoB 2 tool indicated that none of the included peer-reviewed trials were at high risk of bias for the primary outcomes. The main sources of potential bias were the measurement of the outcome (Domain 4) due to the lack of blinding in open-label designs—although this was considered mitigated by the objective nature of the analyzed endpoints—and the selection of the reported result (Domain 5 – D5) in Srivilaithon et al.(25) and Agarwal et al.(30) due to the absence of prospective protocol registration. However, these concerns regarding result selection were deemed mitigated by the high consistency in endpoint reporting observed across all included trials. (eFigure 1-7)

In contrast, the unpublished trial included in the sensitivity analysis (Powers WF, ClinicalTrials.gov NCT03545503, unpublished data) was classified as high risk of bias arising from the randomization process (Domain 1) due to its quasi-randomized allocation method. (eFigure 2).

The certainty of evidence for the primary and secondary outcomes was evaluated using the GRADE framework and is detailed in the Summary of Findings (Table 2). The evidence was rated as high certainty for 28-day all-cause mortality (including the septic patient subgroup), post-intubation hypotension, peri-intubation cardiac arrest, and first-attempt intubation success. The certainty of evidence for hospital resource utilization outcomes, specifically ICU-free days and ventilator-free days, was downgraded to moderate due to imprecision arising from the mathematical approximation of means and standard deviations from the originally reported medians and interquartile ranges.

**Table 2.**
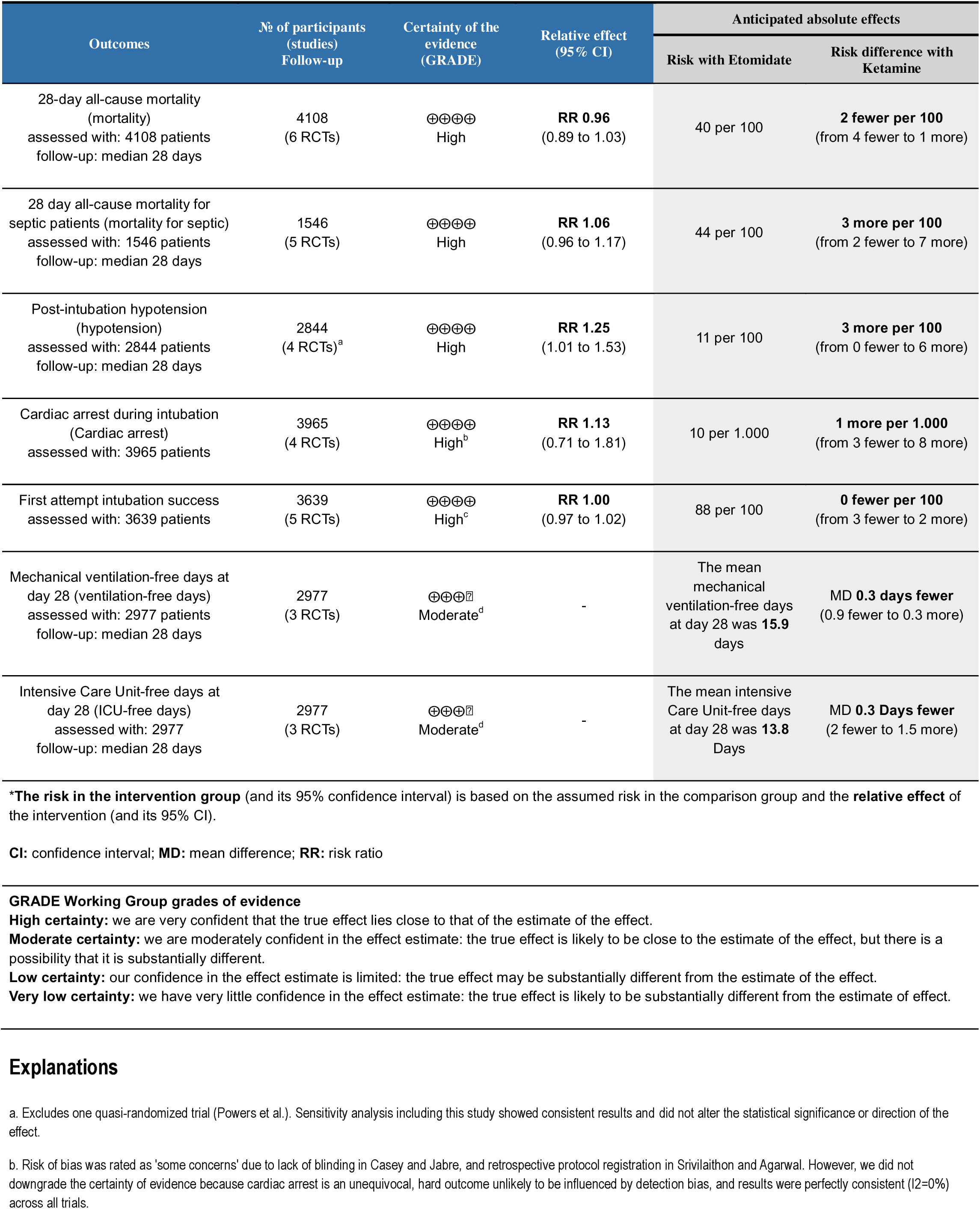
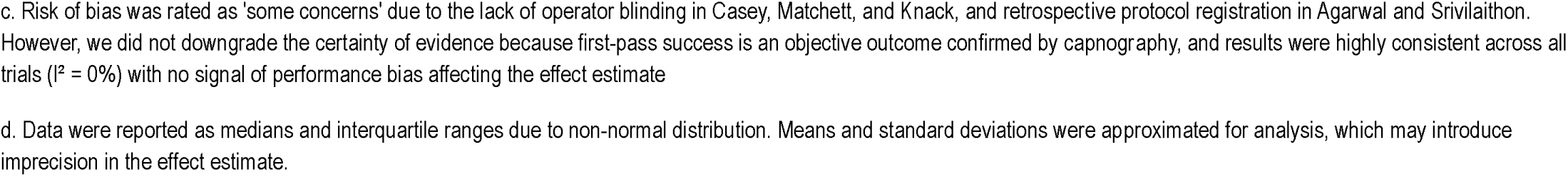
Summary of Findings and Certainty of Evidence (GRADE)

## 4. Discussion

In this systematic review and meta-analysis of six randomized controlled trials involving 4,108 critically ill adults undergoing RSI, we compared the efficacy and safety of etomidate versus ketamine. Our pooled analysis yielded several key findings: (a) there was no statistically significant difference in 28-day all-cause mortality between patients induced with etomidate or ketamine, a finding that persisted in the subgroup analysis of patients with sepsis; (b) ketamine induction was associated with a statistically significant increase in the risk of post-intubation hypotension compared to etomidate; and (c) no significant differences were observed regarding first-pass intubation success, peri-intubation cardiac arrest, or hospital resource utilization (ICU-free and ventilator-free days).

Our findings regarding mortality help resolve a longstanding controversy in critical care medicine. For decades, the use of etomidate has been debated due to its inhibition of 11β-hydroxylase, which causes transient adrenal suppression(6). Previous meta-analyses, often reliant on observational data, suggested this adrenal suppression might translate into increased mortality in septic patients(7,12). However, our results, driven largely by high-quality data from recent large-scale trials, align with the most current literature indicating that single-dose etomidate does not increase mortality(17,18). This suggests that while biochemical adrenal suppression occurs, it does not appear to be clinically relevant to survival outcomes in the context of a single induction dose.

Conversely, our finding regarding hemodynamic stability challenges the traditional dogma that ketamine is the superior agent for preventing hypotension. We observed a statistically significant higher risk of post-intubation hypotension in the ketamine group. While ketamine is a sympathomimetic agent that typically increases blood pressure, it also possesses direct negative inotropic properties(21). In critically ill patients with profound shock who may have depleted catecholamine reserves ("catecholamine washout"), the indirect sympathomimetic support may fail, unmasking the direct myocardial depressant effects(14,22). This "catecholamine depletion" hypothesis is supported by observational data from the National Emergency Airway Registry (NEAR), which also found ketamine associated with higher rates of hypotension and vasopressor requirements compared to etomidate(3,13). Therefore, the assumption that ketamine is universally safer for hemodynamically unstable patients should be reconsidered.

Given the imperfect hemodynamic profile of ketamine and the adrenal suppression associated with etomidate observed in our analysis, future research should explore alternative pharmacological strategies to mitigate these risks. For instance, the theoretical advantage of combining agents with opposing hemodynamic profiles, such as ketamine and propofol (ketofol), has gained traction. The KEEP PACE trial evaluated a reduced-dose ketamine/propofol admixture versus etomidate in critically ill adults and observed no significant differences in post-intubation blood pressure or vasopressor requirements(32). Similarly, the use of adjuvants to offset specific adverse effects—such as co-administering low-dose midazolam with ketamine—represents another viable customized regimen. A prospective trial by Punt et al. comparing etomidate to S-ketamine combined with midazolam for intensive care intubations reported no significant differences in 28-day mortality (38% vs. 39%), duration of norepinephrine support, or length of stay, corroborating that ketamine-based admixtures can yield comparable clinical outcomes(33). Additionally, the individualization of induction agents must heavily weigh patient age and cardiovascular reserve; some evidence suggests that the negative inotropic properties and sympathomimetic failure of ketamine may precipitate myocardial ischemia and severe hypotension particularly in elderly patients(34). Therefore, future large-scale trials should not only focus on single-agent comparisons but also evaluate customized dosing regimens and admixtures in high-risk demographic subgroups.

### 4.1 Limitations

Despite the robustness of these findings, our study has limitations. First, definitions of post-intubation hypotension varied among the included trials, ranging from specific systolic blood pressure thresholds to the initiation of vasopressors. However, statistical heterogeneity for this outcome was zero, suggesting a consistent signal across studies. Second, the largest trials included were open-label regarding the induction agent, which could introduce performance bias; clinicians anticipating hypotension with one agent might have managed periprocedural hemodynamics differently. Third, the dosing of ketamine varied slightly between protocols (1 mg/kg vs. 2 mg/kg), which could influence the degree of hemodynamic fluctuation. Finally, although we reached a large sample size for the primary outcome, rarer adverse events like cardiac arrest may still require even larger sample sizes to detect small differences. Due to the limited number of included studies (N < 10), formal testing for publication bias via funnel plots or Egger’s test could not be performed(19). Consequently, we conducted a sensitivity analysis that incorporated gray literature to address potential bias regarding post-intubation hypotension. The inclusion of these data did not alter the primary findings, thereby reinforcing the validity of our results.

## 5. Conclusion

In this meta-analysis of randomized controlled trials involving critically ill adults, we found no significant difference in 28-day mortality between etomidate and ketamine, including in the subgroup of patients with sepsis. Conversely, ketamine was associated with a statistically significant increase in the risk of post-intubation hypotension compared to etomidate. These findings suggest that while both agents are viable options for RSI, the choice of induction agent should be individualized, with particular caution regarding the potential for hemodynamic instability with ketamine.

## Data Availability

All data produced in the present work are contained in the manuscript

## Acknowledgments (CRediT)

**V. H. A.:** Conceptualization, Methodology, Investigation, Data Curation, Formal Analysis, Project Administration, Writing – Original Draft, and Writing – Review & Editing (participated in all stages of the project as the first author).

**R. P. C.:** Investigation (responsible for screening and study selection) and Methodology (participated in the selection of outcomes of interest).

**B. L. G. F.:** Data Curation (data extraction of outcomes), Formal Analysis, Methodology (helped define methodological issues), and Writing – Review & Editing.

**J. G. S. V.:** Validation (reviewed extracted data for Table 1 and reviewed the PRISMA checklist) and Investigation (full paper availability and retrieval).

**M. A. R. A. N.:** Data Curation (data extraction for Table 1) and Project Administration (contact and partnership establishment with the senior author).

**G. C. R.:** Writing – Review & Editing (final review of the manuscript) and Visualization (preparation of the 280-character summary for social media).

**J. N. Q.:** Supervision and Writing – Review & Editing.

