## Supplementary Material for "Etomidate Versus Ketamine for Emergency Intubation in Critically Ill Patients: An Updated Meta-Analysis and Systematic Review"

^*^Faculdade de Medicina, Universidade de Brasília, Brasília, Brazil.

^†^Departamento de Medicina e Enfermagem, Universidade Federal de Viçosa, Viçosa, Brazil.

^‡^Department of Anesthesiology, Hospital Mãe de Deus, Porto Alegre, Rio Grande do Sul, Brazil, MD

### PRISMA checklist

| **Section and Topic** | **Item #** | **Checklist item** | **Location where item is reported** |
| --- | --- | --- | --- |
| **TITLE** | | |  |
| Title | 1 | Identify the report as a systematic review. | Title, page 1 |
| **ABSTRACT** | | |  |
| Abstract | 2 | See the PRISMA 2020 for Abstracts checklist. | Abstract, page 2 |
| **INTRODUCTION** | | |  |
| Rationale | 3 | Describe the rationale for the review in the context of existing knowledge. | Introduction, page 4 |
| Objectives | 4 | Provide an explicit statement of the objective(s) or question(s) the review addresses. | Introduction, paragraph 4, page 4. |
| **METHODS** | | |  |
| Eligibility criteria | 5 | Specify the inclusion and exclusion criteria for the review and how studies were grouped for the syntheses. | Methods, Eligibility criteria, page 5 |
| Information sources | 6 | Specify all databases, registers, websites, organisations, reference lists and other sources searched or consulted to identify studies. Specify the date when each source was last searched or consulted. | Methods, Search strategy and data extraction, pages 5-6 |
| Search strategy | 7 | Present the full search strategies for all databases, registers and websites, including any filters and limits used. | Supplementary material, pages 4-5 |
| Selection process | 8 | Specify the methods used to decide whether a study met the inclusion criteria of the review, including how many reviewers screened each record and each report retrieved, whether they worked independently, and if applicable, details of automation tools used in the process. | Methods, Search strategy and data extraction, page 5-6 |
| Data collection process | 9 | Specify the methods used to collect data from reports, including how many reviewers collected data from each report, whether they worked independently, any processes for obtaining or confirming data from study investigators, and if applicable, details of automation tools used in the process. | Methods, Search strategy and data extraction, page 5-6 |
| Data items | 10a | List and define all outcomes for which data were sought. Specify whether all results that were compatible with each outcome domain in each study were sought (e.g. for all measures, time points, analyses), and if not, the methods used to decide which results to collect. | Methods, Endpoints (…), Statistical (…), pages 6-7 |
|  | 10b | List and define all other variables for which data were sought (e.g. participant and intervention characteristics, funding sources). Describe any assumptions made about any missing or unclear information. | Results, Study selection (…), paragraph 2, page 8 |
| Study risk of bias assessment | 11 | Specify the methods used to assess risk of bias in the included studies, including details of the tool(s) used, how many reviewers assessed each study and whether they worked independently, and if applicable, details of automation tools used in the process. | Methods, Quality assessment, page 6-7 |
| Effect measures | 12 | Specify for each outcome the effect measure(s) (e.g. risk ratio, mean difference) used in the synthesis or presentation of results. | Methods, Statistical analysis, page 7 |
| Synthesis methods | 13a | Describe the processes used to decide which studies were eligible for each synthesis (e.g. tabulating the study intervention characteristics and comparing against the planned groups for each synthesis (item #5)). | Methods, Eligibility criteria, page 5 |
|  | 13b | Describe any methods required to prepare the data for presentation or synthesis, such as handling of missing summary statistics, or data conversions. | Methods, Statistical analysis, page 7 |
|  | 13c | Describe any methods used to tabulate or visually display results of individual studies and syntheses. | Methods, Statistical analysis, page 7 |
|  | 13d | Describe any methods used to synthesize results and provide a rationale for the choice(s). If meta-analysis was performed, describe the model(s), method(s) to identify the presence and extent of statistical heterogeneity, and software package(s) used. | Methods, Statistical analysis, page 7 |
|  | 13e | Describe any methods used to explore possible causes of heterogeneity among study results (e.g. subgroup analysis, meta-regression). | Results, Subanalyses (…), page 9 |
|  | 13f | Describe any sensitivity analyses conducted to assess robustness of the synthesized results. | Methods, Endpoints (…), page 6 |
| Reporting bias assessment | 14 | Describe any methods used to assess risk of bias due to missing results in a synthesis (arising from reporting biases). | Methods, Quality assessment, page 7 |
| Certainty assessment | 15 | Describe any methods used to assess certainty (or confidence) in the body of evidence for an outcome. | Methods, Quality assessment, page 7 |
| **RESULTS** | | |  |
| Study selection | 16a | Describe the results of the search and selection process, from the number of records identified in the search to the number of studies included in the review, ideally using a flow diagram. | Results, Study selection (…), page 7-8; Figure 1. |
|  | 16b | Cite studies that might appear to meet the inclusion criteria, but which were excluded, and explain why they were excluded. | Supplementary Material, page 5 |
| Study characteristics | 17 | Cite each included study and present its characteristics. | Results, Study selection (…), page 7-8; Table 1 |
| Risk of bias in studies | 18 | Present assessments of risk of bias for each included study. | Results, Quality assessment, page 9-10; Supplementary Material, page 8-11 |
| Results of individual studies | 19 | For all outcomes, present, for each study: (a) summary statistics for each group (where appropriate) and (b) an effect estimate and its precision (e.g. confidence/credible interval), ideally using structured tables or plots. | Results, Pooled analysis (…), page 8-9; Figures 2, 3 and 4 |
| Results of syntheses | 20a | For each synthesis, briefly summarise the characteristics and risk of bias among contributing studies. | Results, Quality assessment, page 9-10 |
|  | 20b | Present results of all statistical syntheses conducted. If meta-analysis was done, present for each the summary estimate and its precision (e.g. confidence/credible interval) and measures of statistical heterogeneity. If comparing groups, describe the direction of the effect. | Results, Pooled analysis (…), page 8-9; Figures 2, 3 and 4 |
|  | 20c | Present results of all investigations of possible causes of heterogeneity among study results. | Results, Pooled analysis (…), page 8-9 |
|  | 20d | Present results of all sensitivity analyses conducted to assess the robustness of the synthesized results. | Results, Subanalyses (…), page 9 |
| Reporting biases | 21 | Present assessments of risk of bias due to missing results (arising from reporting biases) for each synthesis assessed. | Supplementary Material, page 12 |
| Certainty of evidence | 22 | Present assessments of certainty (or confidence) in the body of evidence for each outcome assessed. | Results, Quality assessment, paragraph 3, page 9-10; Table 2 |
| **DISCUSSION** | | |  |
| Discussion | 23a | Provide a general interpretation of the results in the context of other evidence. | Discussion, paragraph 1 and 2, page 10-11 |
|  | 23b | Discuss any limitations of the evidence included in the review. | Discussion, Limitations, page 12-13 |
|  | 23c | Discuss any limitations of the review processes used. | Discussion, Limitations, page 12-13 |
|  | 23d | Discuss implications of the results for practice, policy, and future research. | Discussion, paragraph 4, pages 11-12 |
| **OTHER INFORMATION** | | |  |
| Registration and protocol | 24a | Provide registration information for the review, including register name and registration number, or state that the review was not registered. | Methods, paragraph 1, page 5 |
|  | 24b | Indicate where the review protocol can be accessed, or state that a protocol was not prepared. | Methods, paragraph 1, page 5 |
|  | 24c | Describe and explain any amendments to information provided at registration or in the protocol. | Methods, paragraph 1, page 5 |
| Support | 25 | Describe sources of financial or non-financial support for the review, and the role of the funders or sponsors in the review. | Conflicts of Interest (…), page 1 |
| Competing interests | 26 | Declare any competing interests of review authors. | Conflicts of Interest (…), page 1 |
| Availability of data, code and other materials | 27 | Report which of the following are publicly available and where they can be found: template data collection forms; data extracted from included studies; data used for all analyses; analytic code; any other materials used in the review. | Methods, Data availability, page 7 |

### Search Strategies

- PubMed

(“Critically ill” OR “intubation” OR "Severely ill" OR ICU OR “Critical care” OR Intubation, Intratracheal [MeSH]) AND (Ketamine[MeSH] OR Ketamin* OR Esketamine OR Arketamine OR Ketalar OR Ketaject OR Calypsol OR Ketanest) AND (Etomidate[MeSH] OR Ethomidate OR “Imidazole-5-carboxylate” OR Amidate OR Hypnomidate OR Tomvi)

- Embase

('critically ill patient'/exp OR 'endotracheal intubation'/exp OR 'intensive care unit'/exp OR 'intensive care'/exp OR 'intubation':ti,ab OR 'critically ill':ti,ab OR 'severely ill':ti,ab OR 'icu':ti,ab OR 'critical care':ti,ab) AND ('ketamine'/exp OR 'ketamine':ti,ab OR 'esketamine':ti,ab OR 'arketamine':ti,ab OR 'ketalar':ti,ab OR 'ketaject':ti,ab OR 'calypsol':ti,ab OR 'ketanest':ti,ab) AND ('etomidate'/exp OR 'etomidate':ti,ab OR 'ethomidate':ti,ab OR 'imidazole 5 carboxylate':ti,ab OR 'amidate':ti,ab OR 'hypnomidate':ti,ab OR 'tomvi':ti,ab)

- Cochrane

#1 [mh "Intubation, Intratracheal"]

#2 ("Critically ill" OR "intubation" OR "Severely ill" OR ICU OR "Critical care"):ti,ab,kw

#3 #1 OR #2

#4 [mh Ketamine]

#5 (Ketamin* OR Esketamine OR Arketamine OR Ketalar OR Ketaject OR Calypsol OR Ketanest):ti,ab,kw

#6 #4 OR #5

#7 [mh Etomidate]

#8 (Ethomidate OR "Imidazole-5-carboxylate" OR Amidate OR Hypnomidate OR Tomvi):ti,ab,kw

#9 #7 OR #8

#10 #3 AND #6 AND #9

### Reasons for excluding after full text review

- Study design
  - Williams E, Arthur A, Price B, Banister NJ, Goodloe JM, Thomas SH. Ketamine versus etomidate for use in helicopter emergency medical services endotracheal intubation. Ann Emerg Med. 2012;60(4):S63–4. Located at: Embase. doi:10.1016/j.annemergmed.2012.06.152
    - Observational Study
  - NCT03545503. Evaluating the Hemodynamic Effects of Ketamine Versus Etomidate During Rapid Sequence Intubation. https://clinicaltrials.gov/show/NCT03545503 [Internet]. 2018. Located at: CN-01660046. Disponível em: <https://www.cochranelibrary.com/central/doi/10.1002/central/CN-01660046/full>
    - Excluded from the main analysis due to quasi-randomization method
- Publication Type
  - Letters
    - Davis SC, Hosseinian K. The Effect of Ketamine Versus Etomidate for Rapid Sequence Intubation on Maximum Sequential Organ Failure Assessment Score: A Randomized Clinical Trial. J Emerg Med. janeiro de 2024;66(1):e49. doi:10.1016/j.jemermed.2023.10.036 PubMed PMID: 38342509.
    - Kaufman D. Etomidate versus ketamine for sedation in acutely ill patients. Lancet. 10 de outubro de 2009;374(9697):1240–1; author reply 1241. doi:10.1016/S0140-6736(09)61785-2 PubMed PMID: 19819389
    - Leng Y, Yang Y, Zhou C. Comparing Ketamine and Etomidate for Short-Term Mortality in Rapid Sequence Intubation. Crit Care Med. 1^o^ de maio de 2025;53(5):e1166–7. doi:10.1097/CCM.0000000000006610 PubMed PMID: 40326857
    - Matchett G, Moon TS, Stewart JW, Liang L, Fox PE. Etomidate for endotracheal intubation in sepsis. Author’s reply. Intensive Care Med. 2023;49(3):370–1. Located at: Embase. doi:10.1007/s00134-023-06987-z
    - Mongardon N, Singer M. Etomidate versus ketamine for sedation in acutely ill patients. Lancet. 10 de outubro de 2009;374(9697):1240; author reply 1241. doi:10.1016/S0140-6736(09)61784-0 PubMed PMID: 19819388
  - Protocols
    - DeMasi SC, Imhoff B, Lewis AA, Seitz KP, Driver BE, Gibbs KW, et al. Protocol and Statistical Analysis Plan for a Multicenter Randomized Trial of Ketamine vs Etomidate for Emergency Tracheal Intubation. CHEST Crit Care. setembro de 2025;3(3):100177. doi:10.1016/j.chstcc.2025.100177 PubMed PMID: 41472917; PubMed Central PMCID: PMC12747562.
    - Ketamine Versus Etomidate for Sedation of Emergency Department Patients During Rapid Sequence Intubation. clinicaltrials.gov [Internet]. 2013. Disponível em: <https://www.embase.com/search/results?subaction=viewrecord&id=LNCT01823328&from=export>
  - Notes
    - Ecker-Schlipf B. Endotracheal intubation: Ketamine as alternative to etomidate. Krankenhauspharmazie. 2010;31(6):287–8. Located at: Embase
  - Editorial
    - Leibowitz AB. Ketamine Versus Etomidate for Endotracheal Intubation of Critically Ill Patients. Crit Care Med. 1^o^ de fevereiro de 2025;53(2):e504–7. doi:10.1097/CCM.0000000000006536 PubMed PMID: 39982188.
  - Interim analysis
    - Nakajima S, Taylor K, Hall Zimmerman L, Collopy K, Fales C, Powers W. Hemodynamic effects of ketamine versus etomidate during rapid sequence intubation in an ED. Crit Care Med [Internet]. 2019;47(1). Located at: Embase. Disponível em: <https://www.embase.com/search/results?subaction=viewrecord&id=L629628303&from=export>
      - Conference abstract with preliminary results of Knack et. al.
  - Commentaries
    - Trikha A. Acutely ill patients: rapid sequence intubation with etomidate or ketamine. Natl Med J India. dezembro de 2009;22(6):308–9. PubMed PMID: 20384021.
    - Ketamine Versus Etomidate During Rapid Sequence Intubation: Consequences on Hospital Morbidity. clinicaltrials.gov [Internet]. 2007. Disponível em: <https://www.embase.com/search/results?subaction=viewrecord&id=LNCT00440102&from=export>
  - Duplicated trial registries
    - Etomidate Versus Ketamine for Emergency Endotracheal Intubation: a Prospective Randomized Clinical Trial. clinicaltrials.gov [Internet]. 2015. Disponível em: <https://www.embase.com/search/results?subaction=viewrecord&id=LNCT02643381&from=export>
    - NCT02643381. Etomidate Versus Ketamine for Emergency Endotracheal Intubation: a Prospective Randomized Clinical Trial. https://clinicaltrials.gov/show/NCT02643381 [Internet]. 2015. Located at: CN-01554782. Disponível em: <https://www.cochranelibrary.com/central/doi/10.1002/central/CN-01554782/full>
- Review
  - Bruder EA, Ball IM, Ridi S, Pickett W, Hohl C. Single induction dose of etomidate versus other induction agents for endotracheal intubation in critically ill patients. Cochrane Database Syst Rev. 2015;2017(6). Located at: Embase. doi:10.1002/14651858.CD010225.pub2
  - Long B, Gottlieb M. Ketamine versus etomidate for induction of intubation in critically ill patients. Acad Emerg Med. setembro de 2024;31(9):937–8. doi:10.1111/acem.14941 PubMed PMID: 38783635
- Interrupted Study
  - NCT04120870. Comparison of Ketamine and Etomidate During Rapid Sequence Intubation in Trauma Patients. https://clinicaltrials.gov/show/NCT04120870 [Internet]. 2019. Located at: CN-01992704. Disponível em: <https://www.cochranelibrary.com/central/doi/10.1002/central/CN-01992704/full>
- Long term report
  - Effect of Ketamine and Etomidate During Rapid Sequence Intubation on Long- Term Outcomes (Long-Term Outcomes of the RSI Trial). clinicaltrials.gov [Internet]. 2023. Disponível em: <https://www.embase.com/search/results?subaction=viewrecord&id=LNCT06179485&from=export>

### RoB-2 results report

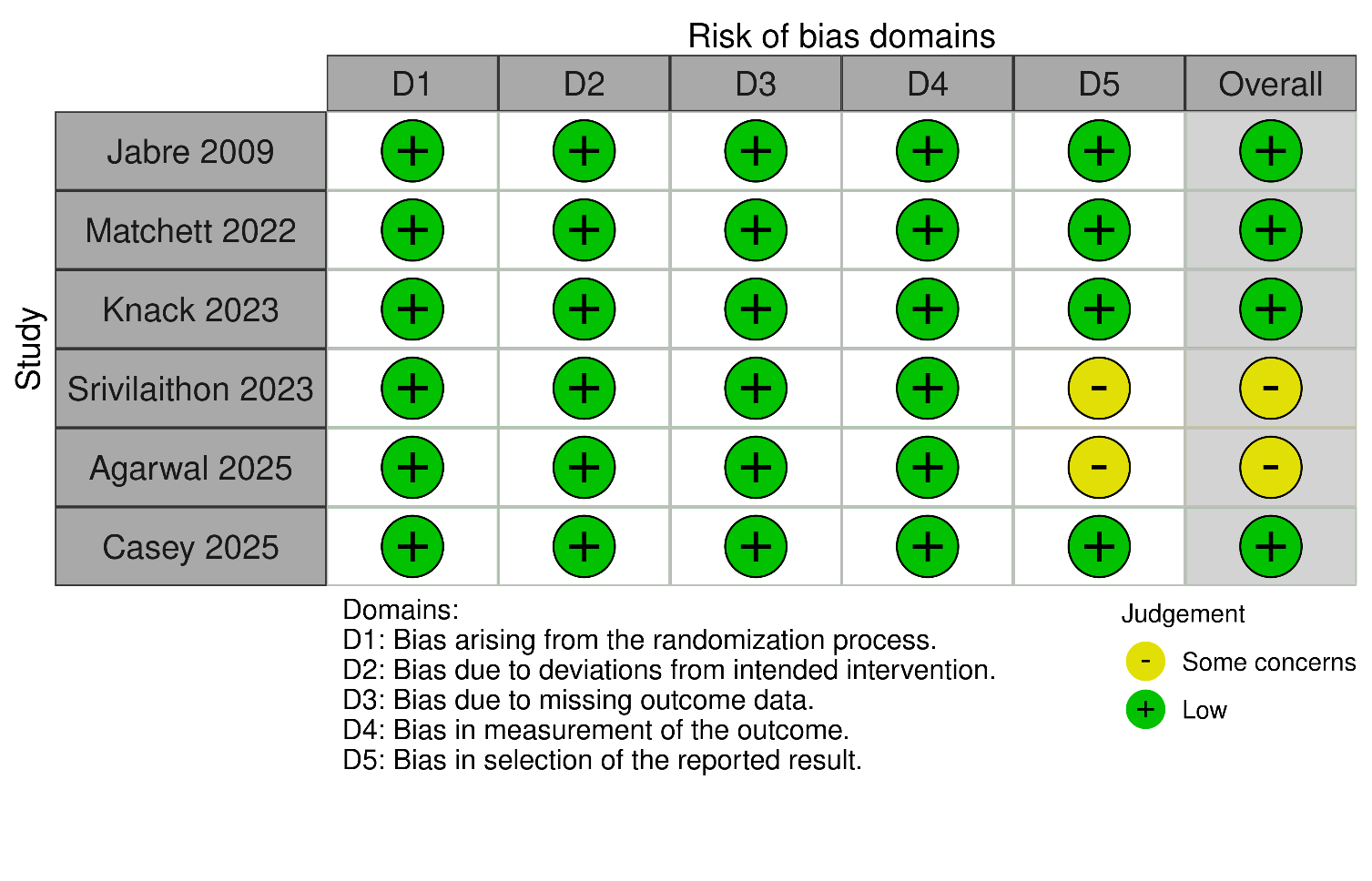

#### eFigure 1. Risk of bias assessment for the 28-day mortality outcome.

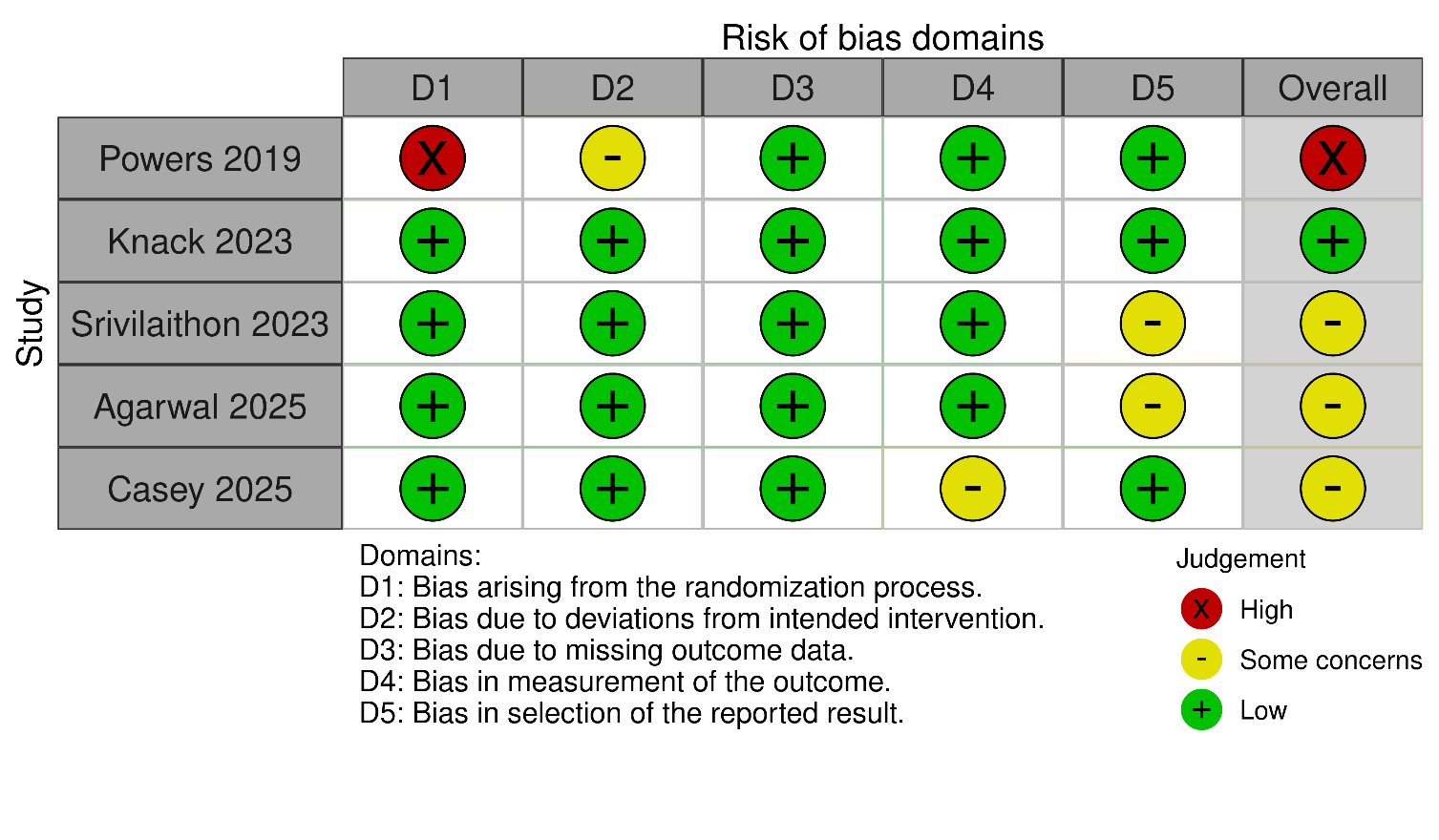

#### eFigure 2. Risk of bias assessment for the post-intubation hypotension outcome, including the quasi-RCT Powers et. al.

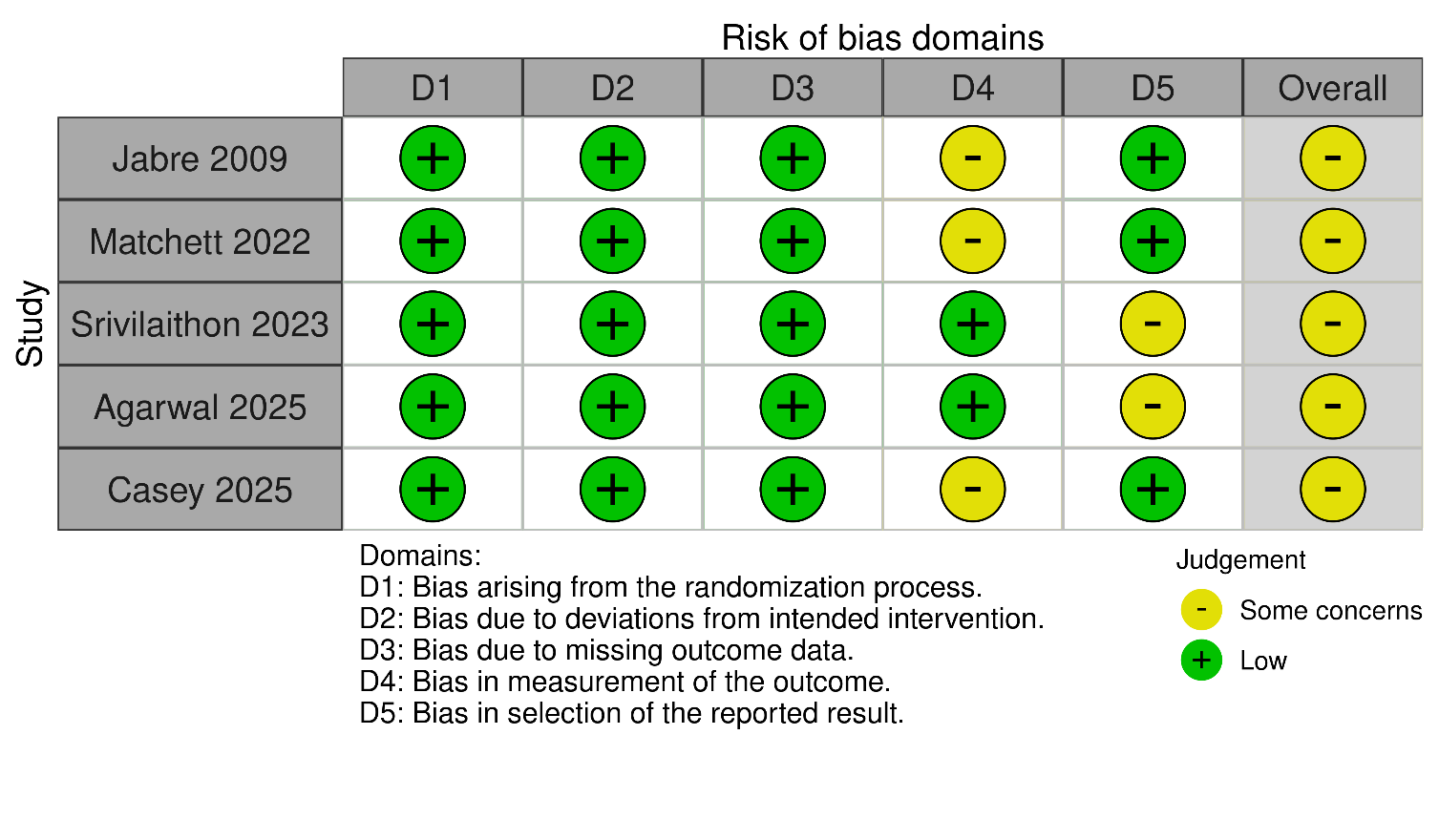

#### eFigure 3. Risk of bias assessment for the peri-intubation cardiac arrest outcome.

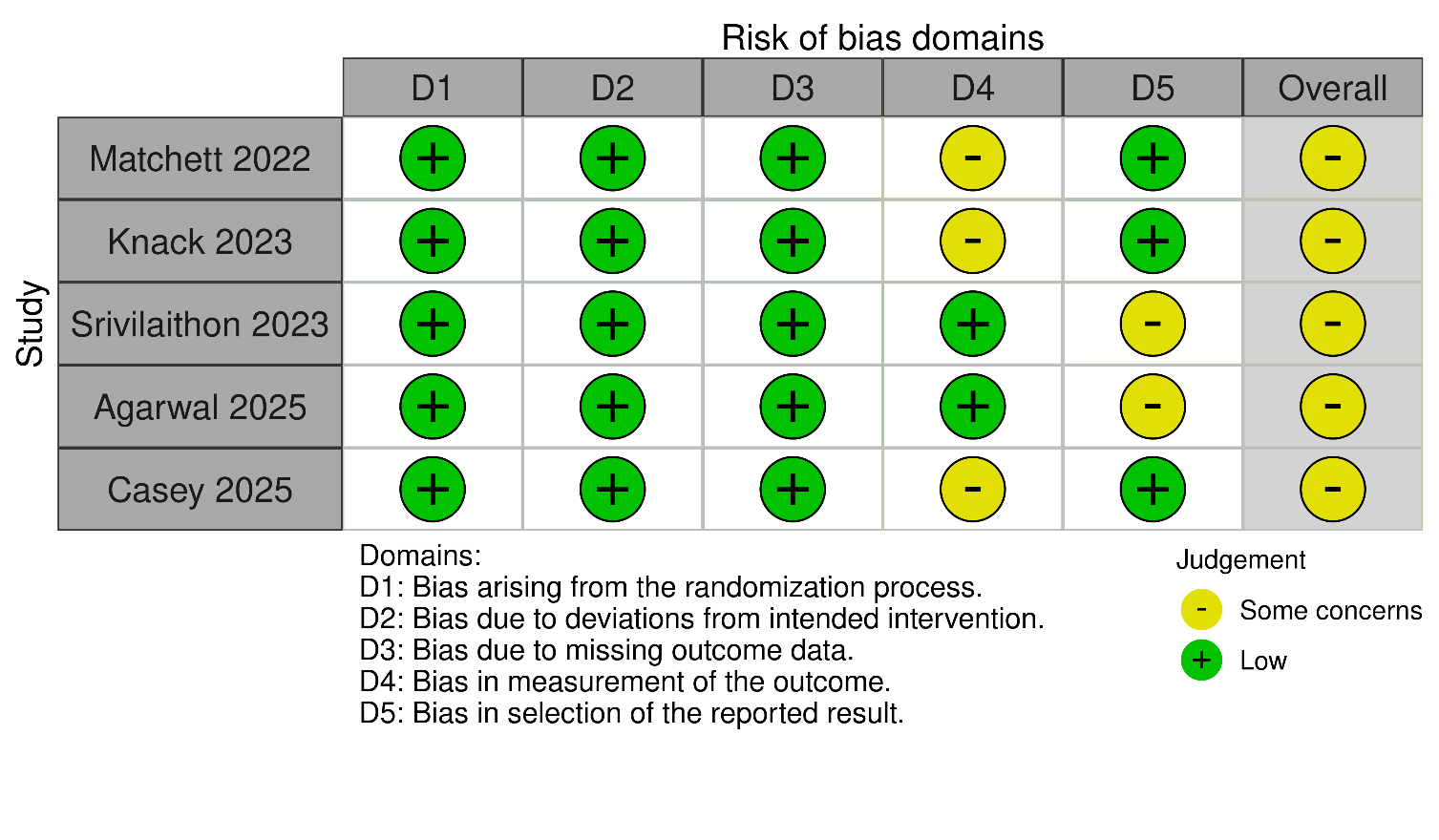

#### eFigure 4. Risk of bias assessment for the first attempt intubation success outcome.

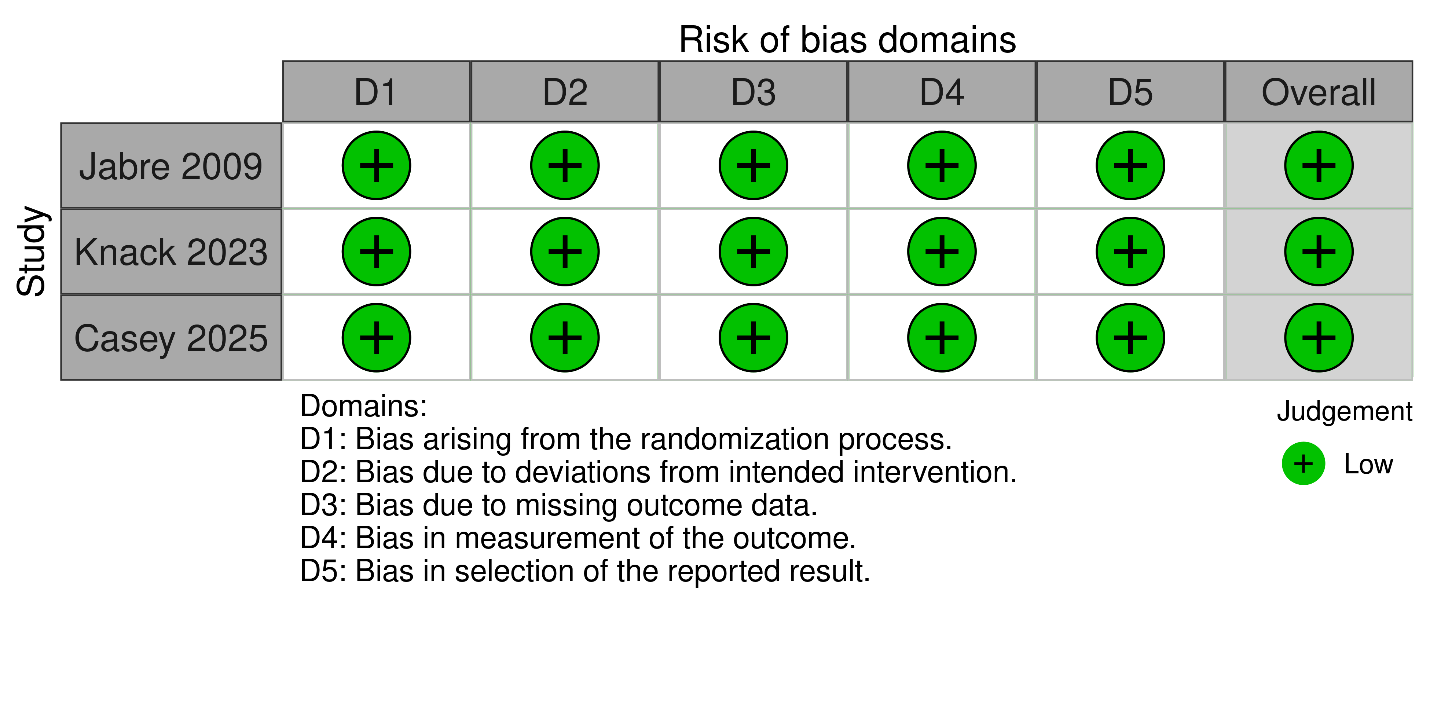

#### eFigure 5. Risk of bias assessment for the ICU-free days outcome.

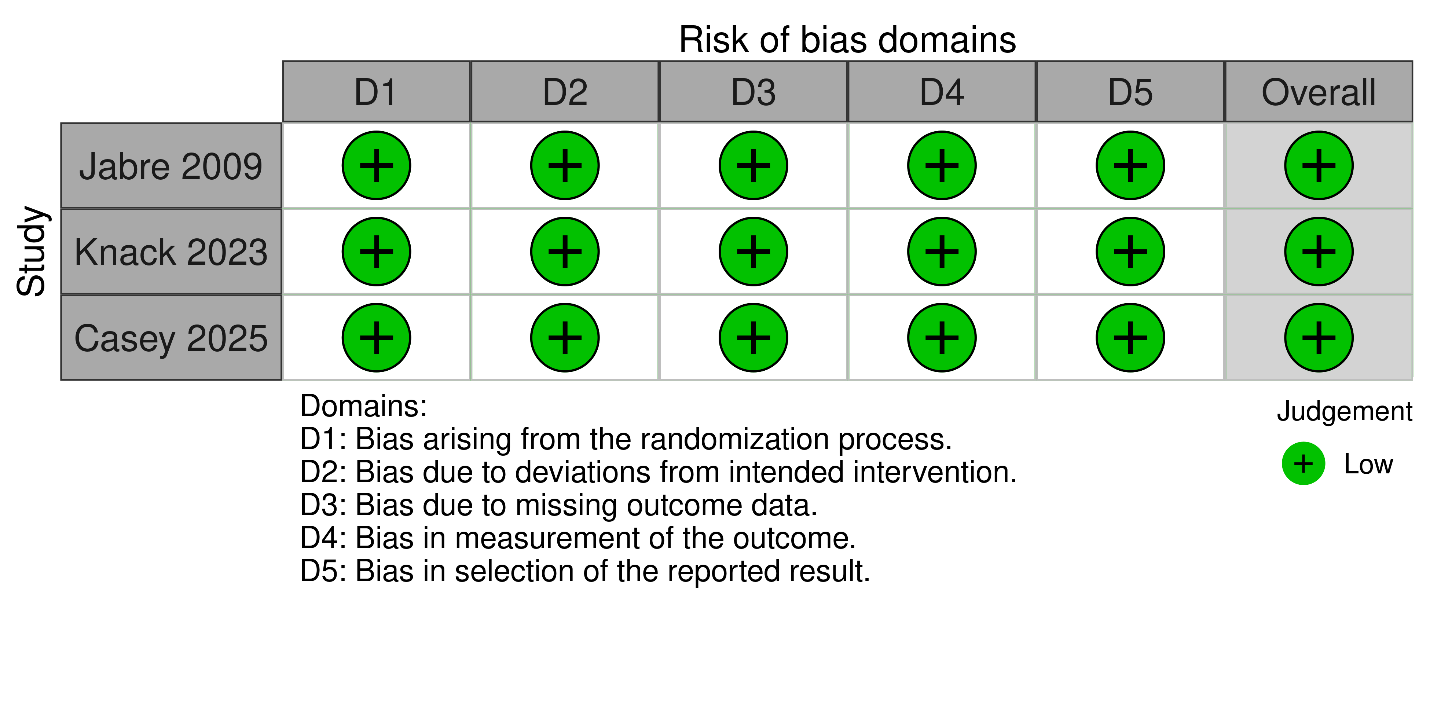

#### eFigure 6. Risk of bias assessment for the mechanical ventilator-free days outcome.

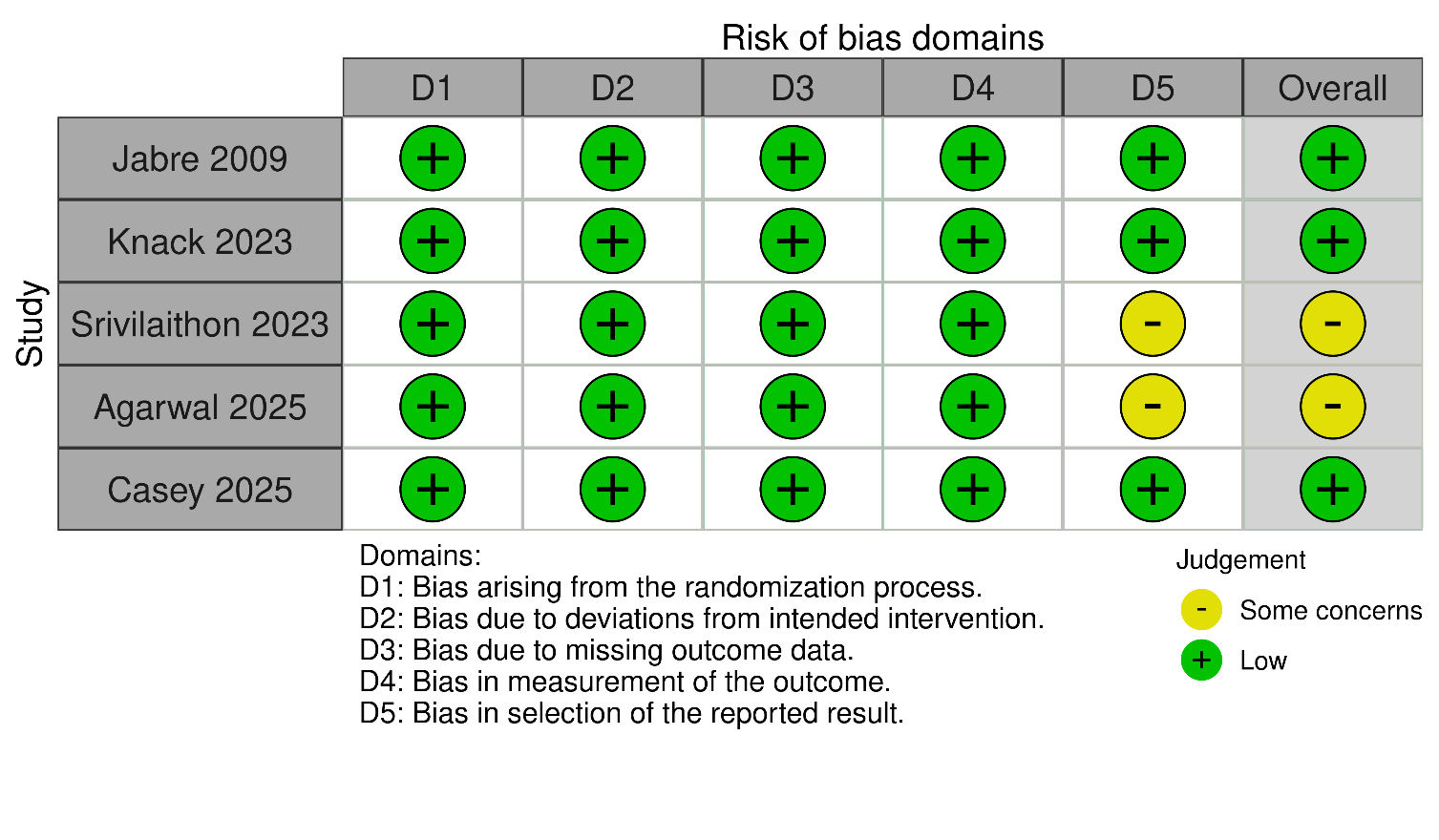

#### eFigure 7. Risk of bias assessment for the 28-day mortality outcome of the septic subgroup.

### Sensitivity analysis

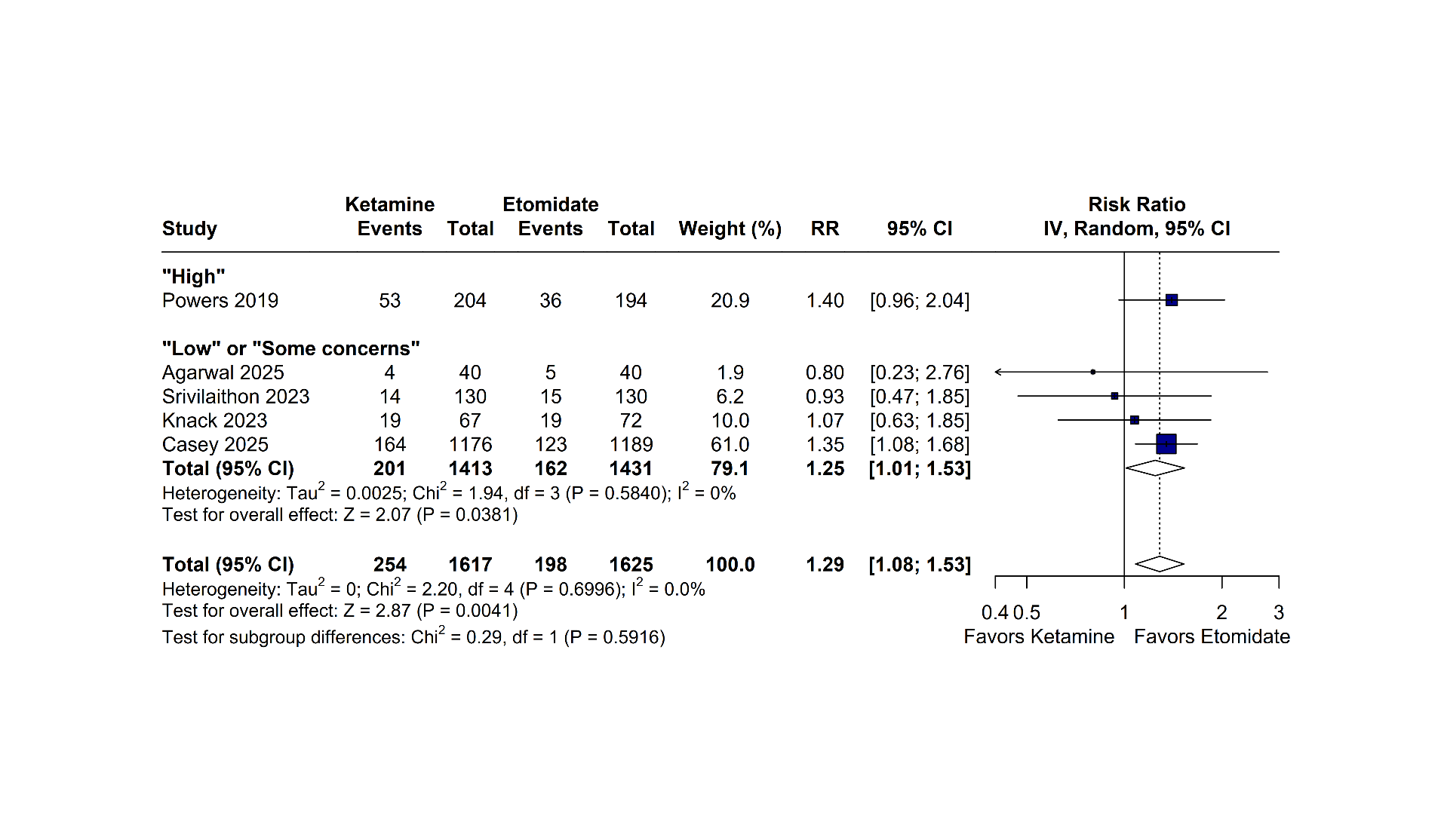

#### eFigure 8. Sensitivity analysis of post-intubation hypotension.

Forest plots comparing the pooled effect estimates of rapid sequence intubation using ketamine versus etomidate, stratified by risk of bias to assess the impact of including a quasi-randomized trial (Powers 2019). CI = Confidence Interval; IV = Inverse Variance; RR = Risk Ratio.

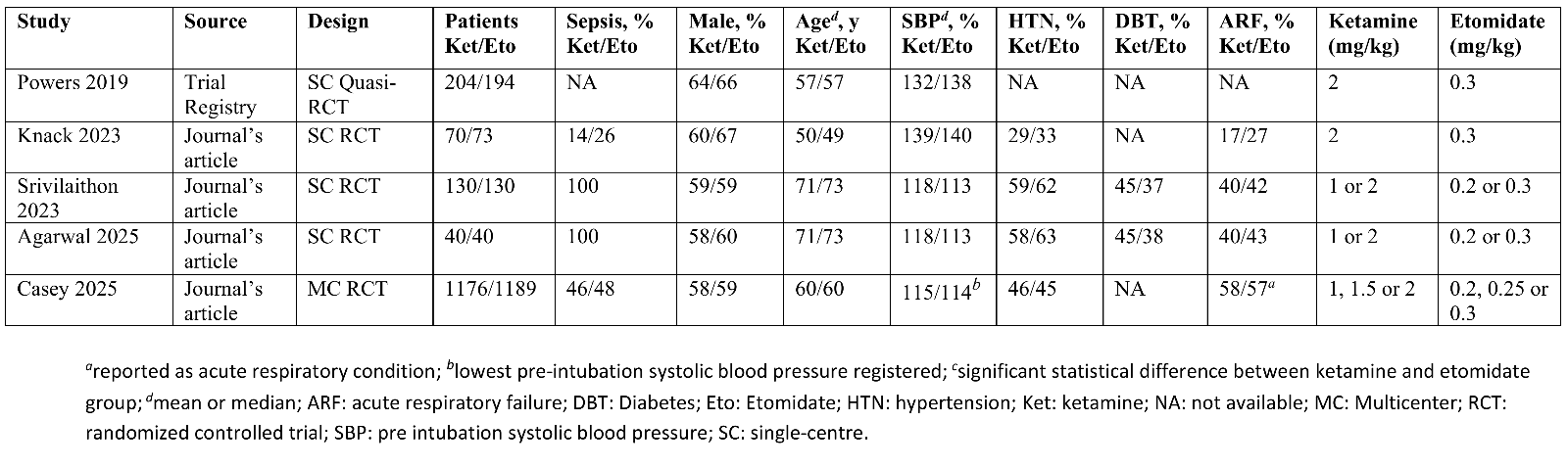

#### eTable 1. Baseline characteristics of included studies in post-induction hypotension sensitivity analysis.
